# Evaluation the angle of posterosuperior horn of normal fetal Sylvian fissure and its morphological changes by trans-cerebellar section

**DOI:** 10.1101/2021.05.07.21256817

**Authors:** Xu Pingping, Zhang Dirong, Shi Yu, Kong Fengbei, Yao Chunxiao, She Ying, Wu Guoru

## Abstract

**Objective:** On the basis of retrospectively analysis the trans-cerebellar section showing the Sylvian fissure of normal fetus is better than trans-thalamic section in middle and late trimester, we prospectively studied the morphological changes of the Sylvian fissure of normal fetus on the trans-cerebellar section in order to provide valuable information for the diagnosis of fetal cerebral cortical dysplasia.

**Methods:** A prospective cross-sectional study was conducted on 845 normal fetuses at 21 to 32 weeks of gestation from January 2019 to September 2020. The angle of the posterosuperior horn of the Sylvian fissure was measured. Based on the angle, the morphology of the Sylvian fissure was divided into four shapes included 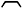 shape (Angle >90°), П shape (Angle ≈90°), π shape (Angle <90°) and *工* shape (Angle ≈0°).

**Results:** The angle of the posterosuperior horn of the Sylvian fissure was negatively correlated with gestational age which correlation coefficient was 0.966 (P < 0.001). Taking gestational age as the independent variable and the angle of posterosuperior horn of the Sylvian fissure as the dependent variable, it showed that there was a linear relationship between the gestational age and the angle of posterosuperior horn of the Sylvian fissure. We got a simple correlation formula that the angle of posterosuperior horn of the Sylvian fissure (°) =140-13×(gestational weeks-21). It was found that the morphological changes of the Sylvian fissure were related to the gestational age. The morphology of the Sylvian fissure was 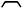-shaped at 21 to 24 weeks of gestation, П-shaped at 25 to 26 weeks of gestation, π-shaped at 26 to 30 weeks of gestation, and the Sylvian fissure was almost closed and appeared *工* shape after 31 to 32 weeks of gestation.

**Conclusion:** This study preliminarily elucidates that the morphological changes of the posterosuperior horn of the Sylvian fissure of normal fetuses at 21 to 32 weeks of gestation through the trans-cerebellar section, which could provide valuable information for the evaluation of the normal development of the Sylvian fissure and the prenatal diagnosis of cerebral cortical hypoplasia of fetuses.

## INTRODUCTION

Cerebral cortical developmental malformation is difficult to diagnose by prenatal ultrasound^1.^ However, it seriously affects the quality life of children and could lead to severe intellectual retardation and intractable epilepsy^2^. Deformity of fetal cerebral cortex is usually manifested as abnormal sulcus development, which can be specifically manifested as delayed sulcus development, non-development of sulcus and overdevelopment of sulcus^3^. The delay of sulci development means that sulci development is more than 3 weeks later than that of normal healthy fetus of the same age^4^. The Sylvian fissure, located on the lateral side of the cerebral hemisphere, is one of the main cerebral sulcus. The Sylvian fissure is the earliest one appeared in the development of all the sulci^5^, and its development is representative. Therefore, the development law of the Sylvian fissure can be further indicated by studying the development law of the cerebral cortex. Some studies have reported the correlation between the depth and width of the Sylvian fissure with gestational age in normal fetuses, but there are few studies on the change of the angle and morphology of the Sylvian fissure. Besides, all the studies have quantitatively evaluated the depth and width of the Sylvian fissure on the trans-thalamic section or the customized section. In our previous retrospective study, we have found that the Sylvian fissure was shown clearer on trans-cerebellar section than trans-thalamic section. Therefore, the aim of this study was to establish the normal reference range of the angle of posterosuperior horn of the Sylvian fissure of normal fetuses in middle and late trimester by the trans-cerebellar section, and to evaluate the change rule of the morphology of the Sylvian fissure, so as to provide valuable information for the evaluation of the normal development of fetal Sylvian fissure and the suspicious diagnosis of fetal cerebral cortical dysplasia by prenatal ultrasound.

## METHODS

This was a prospective cross-sectional study. Normal fetuses undergoing routine prenatal ultrasonography in our hospital from January 2019 to September 2020 were selected as subjects. To exclude abnormal fetuses and ensure the relative accuracy of gestational age, the inclusion criteria were as follows: the date of the last menstruation was clear or the gestational week was corrected by crown-lump length;21 to 32 weeks of gestation; low risk pregnancy including pregnant women without diabetes, hypertension and other pregnancy risk factors. Exclusion criteria were as follows: fetuses with known malformations or chromosomal abnormalities; the large difference between the biological gestational age and the actual gestational age of the fetuses; multiple pregnancies; nervous system abnormality occurs within 6 months after delivery.

This study was approved by the Peking University Shenzhen Hospital and Health Service Human Research Ethics Committee. All pregnant women gave informed consent to this study. The pregnant women ranged in age from 20 to 46 years old, with an average age of 27 years old. The gestational age ranged from 21 to 32 weeks, with an average gestational age of 27 weeks.

The ultrasound machine used in this study was a Voluson E8 or E10 (GE Healthcare Ultrasound) with a 4-8MHz transabdominal 2D transducer. In each examination, the biparietal diameter, head circumference, abdominal circumference, femur and other parameters of the fetus were measured to comprehensively evaluate the biological gestational age of the fetus, and then the screening of the structural malformation of each fetal system was conducted.

When detailed screening of fetal central nervous system structures, the angle of posterosuperior horn of the Sylvian fissure of the fetus was measured by trans-cerebellar section. Based on the angle, the morphology of the Sylvian fissure was divided into four shapesincluded 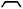 shape (Angle >90°), П shape (Angle ≈90°), π shape (Angle <90°) and *工* shape (Angle ≈0°). Standard trans-cerebellar section^6^: the probe clearly showed the midline of the brain, cavity of septum pellucidum, thalamus, bilateral cerebellar hemispheres and the vermis of the cerebellum, posterior fossa and Sylvian fissure. The measurement method was shown in Figure 1. The angle of posterosuperior horn of the Sylvian fissure was formed between the top and rear edge of the Sylvian fissure. Each patient was measured by one experienced sonographer, and each was measured three times. When the fetus had poor image acquisition due to positional limitations, the pregnant woman was instructed to perform the scan after moderate activity.

**Fig. 1.**
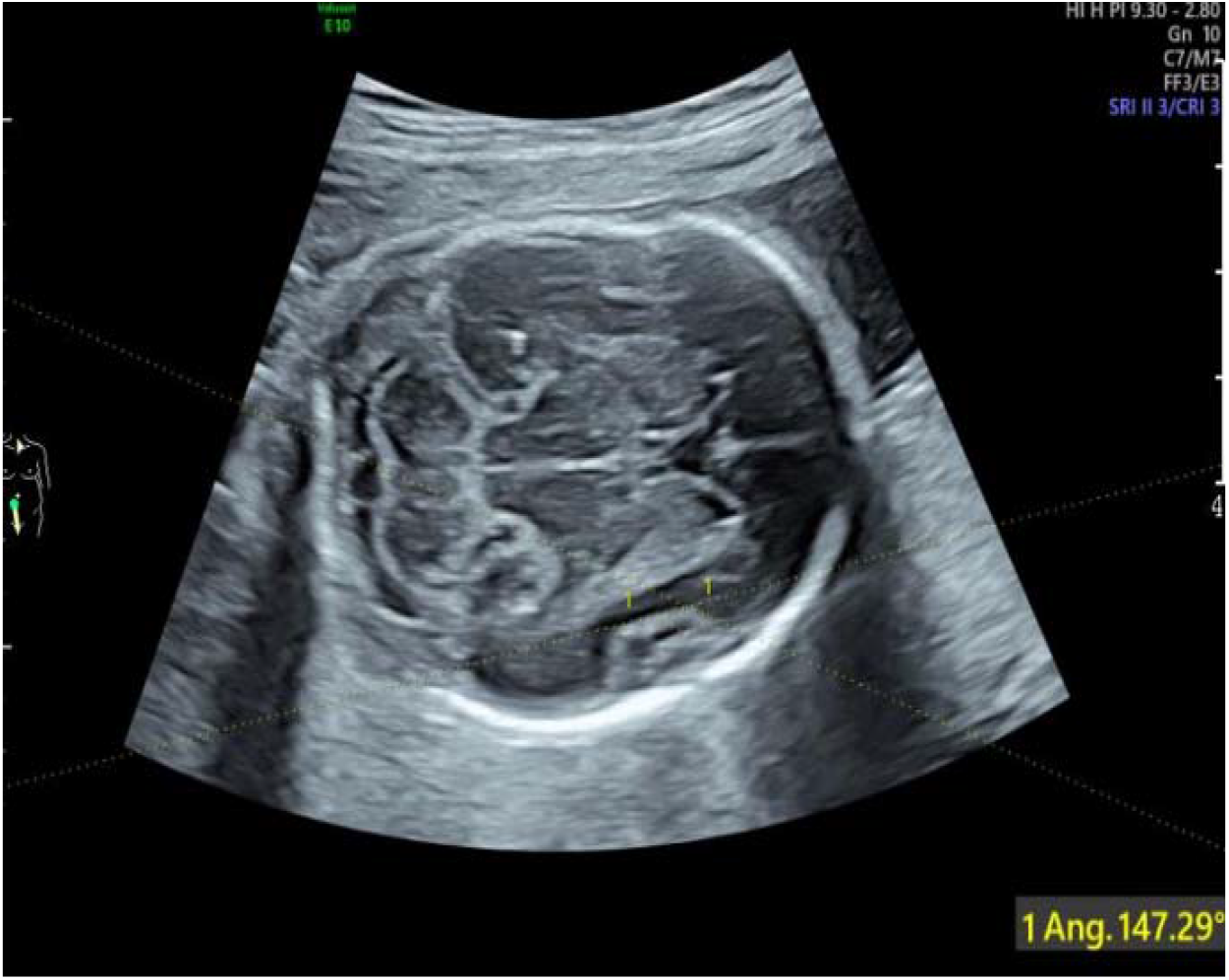
Measurement of the angle of posterosuperior horn of the Sylvian fissure at 23 weeks of gestation

SPSS 26.0 statistical software was used to analyze and process the data. The main steps were as follows:(i) All measured data were counted and divided into 12 groups according to gestational age. The gestational age sample size of each group was counted, the mean value and standard deviation of each parameter in each group 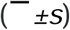 were calculated;(ii)the normality of each parameter value was tested by the Anderson-Darling test; (iii) for data that conformed to a normal distribution, the 95% reference range was calculated using the two-sided boundary value normal distribution method; for data with a skewed distribution, the normal reference range was calculated using the two-sided boundary value percentile method. The scatter plot of the relationship between the angle of posterosuperior horn of the Sylvian fissure of the fetus and the week of gestation was drawn, and a linear regression equation was established by linear regression analysis, with *p*<0.05 being statistically significant.

## RESULTS

A total of 845 subjects were included in the study. The sample data of the angle of posterosuperior horn of the Sylvian fissure of the fetus at different gestational weeks were consistent with or approximately normal distribution. The mean value, standard deviation and 95% reference range of each parameter were calculated at different gestational weeks, and the bilateral boundary value was taken. The data were shown in Table 1.

**Table 1.** The mean value, standard deviation 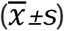 and 95% reference range of the angle of posterosuperior horn of the Sylvian fissure of fetus at different gestational weeks

| Gestational weeks | Numbers | angle of posterosuperior horn of the Sylvian fissure |  |
| --- | --- | --- | --- |
| | | ($\bar{x} \pm s$) | 95% reference range |
|  |  | range |  |
| 21 | 65 | 129.91±8.45 | 113.97~142.99 |
| 22 | 70 | 122.55±10.36 | 101.33~140.28 |
| 23 | 91 | 119.47±12.32 | 93.03~139.17 |
| 24 | 75 | 104.85±12.17 | 83.98~123.13 |
| 25 | 68 | 93.61±7.18 | 81.78~110.69 |
| 26 | 78 | 76.53±10.54 | 57.42~93.76 |
| 27 | 75 | 69.27±12.52 | 46.78~92.95 |
| 28 | 70 | 58.55±12.81 | 36.24~82.06 |
| 29 | 71 | 37.06±8.30 | 24.02~52.82 |
| 30 | 67 | 16.63±12.63 | 0.00~34.00 |
| 31 | 60 | 9.32±8.79 | 0.00~27.85 |
| 32 | 55 | 1.82±4.92 | 0.00~14.03 |

A scatterplot of the relationship between the angle of posterosuperior horn of the Sylvian fissure of the fetus and gestational weeks was drawn (Figure 2). Correlation analysis showed that the angle of posterosuperior horn of the Sylvian fissure of the fetus was negatively correlated with gestational weeks, with a correlation coefficient 0.966 (P < 0.001). After 31 weeks of gestation, the angle of posterosuperior horn of the Sylvian fissure was nearly 0° and the Sylvian fissure was closed morphologically.

**Fig. 2.**
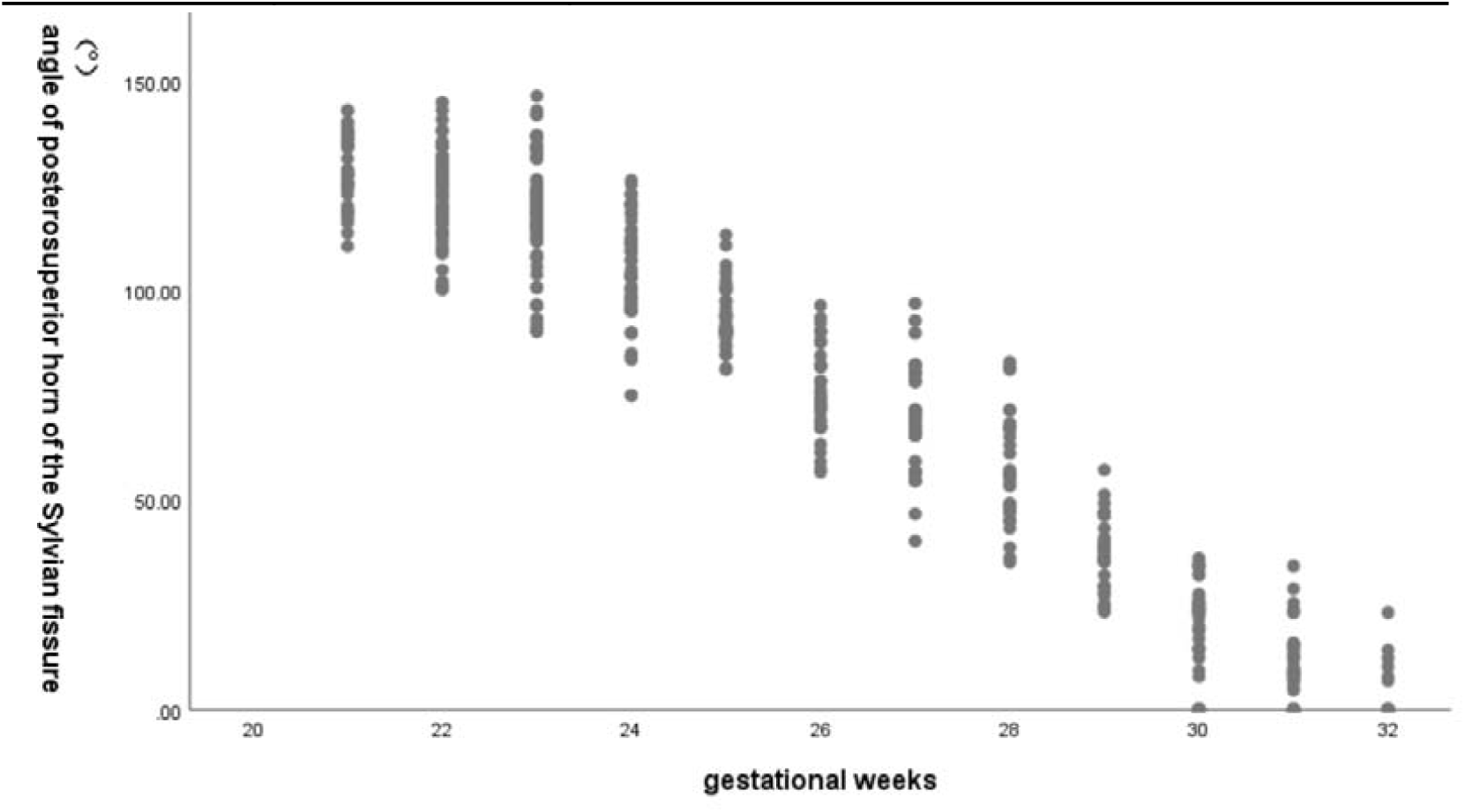
Scatterplot of the relationship between the angle of posterosuperior horn of the Sylvian fissure and gestational weeks

Linear regression analysis was conducted with gestational weeks as the independent variable and the angle of posterosuperior horn of the Sylvian fissure as the dependent variable. The result showed that there was a linear regression relationship between the gestational weeks of the fetus and the angle of posterosuperior horn of the Sylvian fissure, and the obtained linear regression equation was the angle of posterosuperior horn of the Sylvian fissure (°)=140.178-12.715×(gestational weeks-21).The equation was simplified as follows: the angle of posterosuperior horn of the Sylvian fissure (°)=140-13× (gestational weeks-21). It was found that the morphological changes of the Sylvian fissure were related to the gestational age. Based on the angle, the morphology of the Sylvian fissure was divided into four shapes included 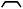 shape (Angle >90°), П shape (Angle ≈90°), π shape (Angle <90°) and *工* shape (Angle ≈0°).The morphology of the Sylvian fissure was 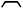-shaped at 21 to 24 weeks of gestation, П-shaped at 25 to 26 weeks of gestation, π-shaped at 26 to 30 weeks of gestation, and the Sylvian fissure was almost closed and appeared after *工* shape 31 to 32 weeks of gestation(Figure 3).

**Fig. 3.**
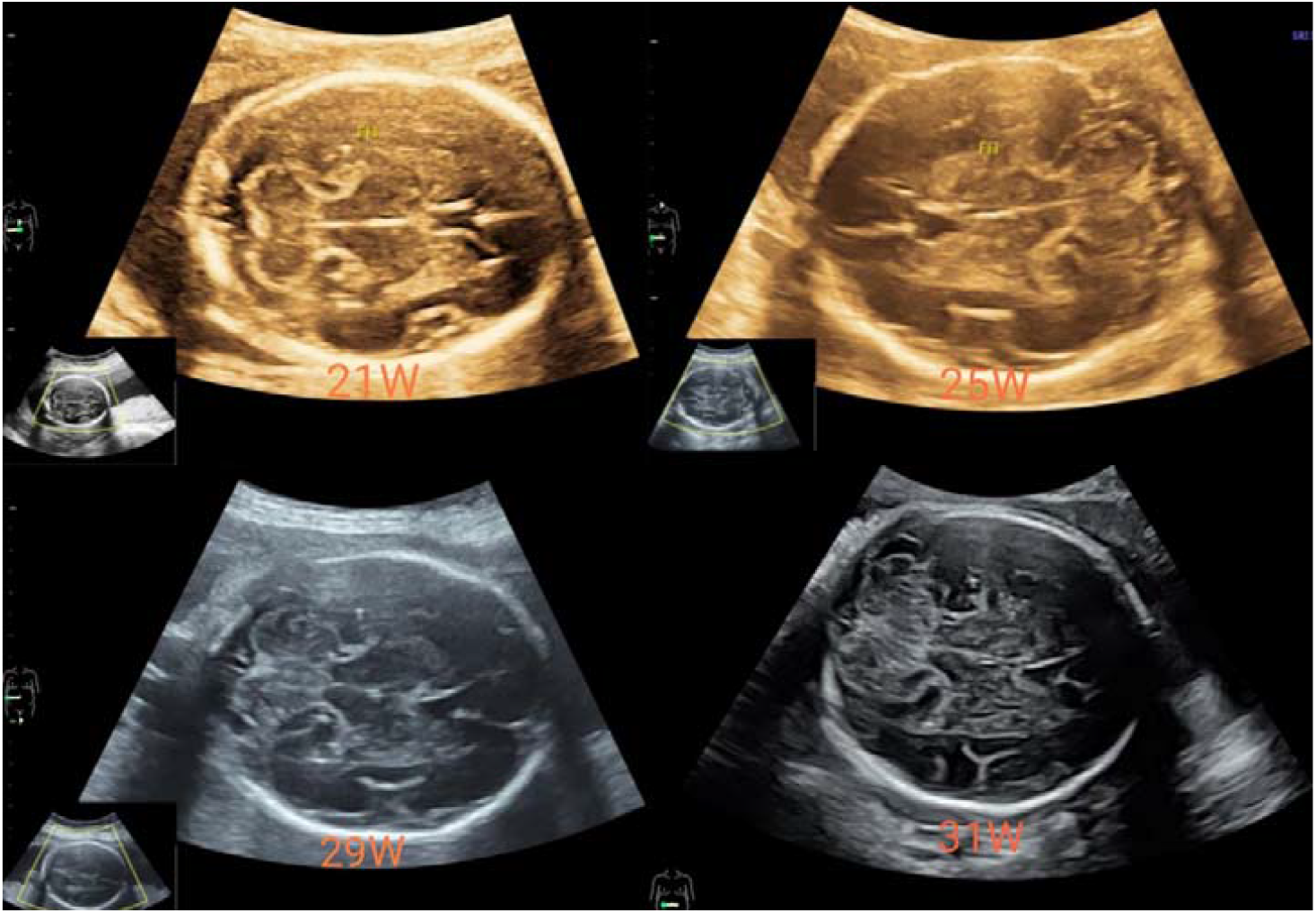
four shapes of the morphology of the Sylvian fissure included 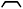 shape (Angle >90°), П shape (Angle ≈90°), π shape (Angle <90°) and *工* shape (Angle ≈0°)

## DISCUSSION

Human brain development is extremely complex, but it follows strict developmental rules. Some scholars consider that cerebral sulci can be used as a sign of fetal cranial development^7^. The developmental process of cerebral sulci can be used as a standard to evaluate the gestational age. By observing the development of cerebral sulci to further evaluate the development of the whole fetal brain is of great significance to the evaluation of fetal brain development and the diagnosis of diseases. The brain develops rapidly and has obvious morphological changes. In appearance, it can be seen that the brain develops from a tubular structure to a surface covered with furrows, and various branches of the furrows will appear in the process of forming sulci^8-10^.

The Sylvian fissure, located on the lateral side of the cerebral hemisphere, is one of the major cerebral sulcus and the earliest one sulci to appear. It separates the upper frontal and parietal lobes from the lower temporal lobes. The development of the Sylvian fissure also indirectly reflects the development speed of the frontal, parietal and temporal lobes^11^. Prenatal ultrasound can clearly show the Sylvian fissure, so it can be used to evaluate the development of the Sylvian fissure. Current studies have assessed the development of fetal Sylvian fissure^12-13^, but trans-thalamic sections were mainly used to study the depth and width of Sylvian fissure. In our previous retrospective study, we have found that the trans-cerebellar transverse section showed the Sylvian fissure clearer than the trans-thalamic section in each gestational week, which indicated that most of the developmental parameters related to the Sylvian fissure can be measured in the trans-cerebellar section, and the measurement results will be more repeatable. The overall clarity of the Sylvian fissure shown on the trans-thalamic section is low, which indicated that in most cases, adjustments are needed on this standard section to measure the parameters related to the Sylvian fissure, which may affect the repeatability of the measurement results.

In this study, normal fetuses from 21 to 32 weeks of gestation were selected as the research object, and trans-cerebellar section was selected and the data were successfully measured with a rate of 95.6%. The angle of the posterosuperior horn of the Sylvian fissure was negatively correlated with gestational weeks at 21 to 32 weeks which correlation coefficient was 0.966 (P < 0.001). Taking gestational weeks as the independent variable and the angle of posterosuperior horn of the Sylvian fissure as the dependent variable, it showed that there was a linear relationship between the gestational weeks and the angle of posterosuperior horn of the Sylvian fissure. We got a simple correlation formula that the angle of posterosuperior horn of the Sylvian fissure (°) =140-13× (gestational weeks -21). Based on the angle, the morphology of the Sylvian fissure was divided into four shapes included 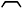 shape (Angle >90°), П shape (Angle ≈90°), π shape (Angle <90°) and *工* shape (Angle ≈0°). It was found that the morphological changes of the Sylvian fissure were related to the gestational weeks. The morphology of the Sylvian fissure was 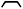-shaped at 21 to 24 weeks of gestation, П-shaped at 25 to 26 weeks of gestation, π-shaped at 26 to 30 weeks of gestation, and the Sylvian fissure was almost closed and appeared after *工* shape 31 to 32 weeks of gestation. If the angle or morphological changes of the Sylvian fissure were found significantly behind the gestational week during prenatal ultrasound examination (as shown in Figure 4 and Figure 5). We should be alert to the possibility of delayed development of cerebral sulci in the fetus. MRI examination of the brain of the fetus confirmed cortical dysplasia.

**Fig. 4.**
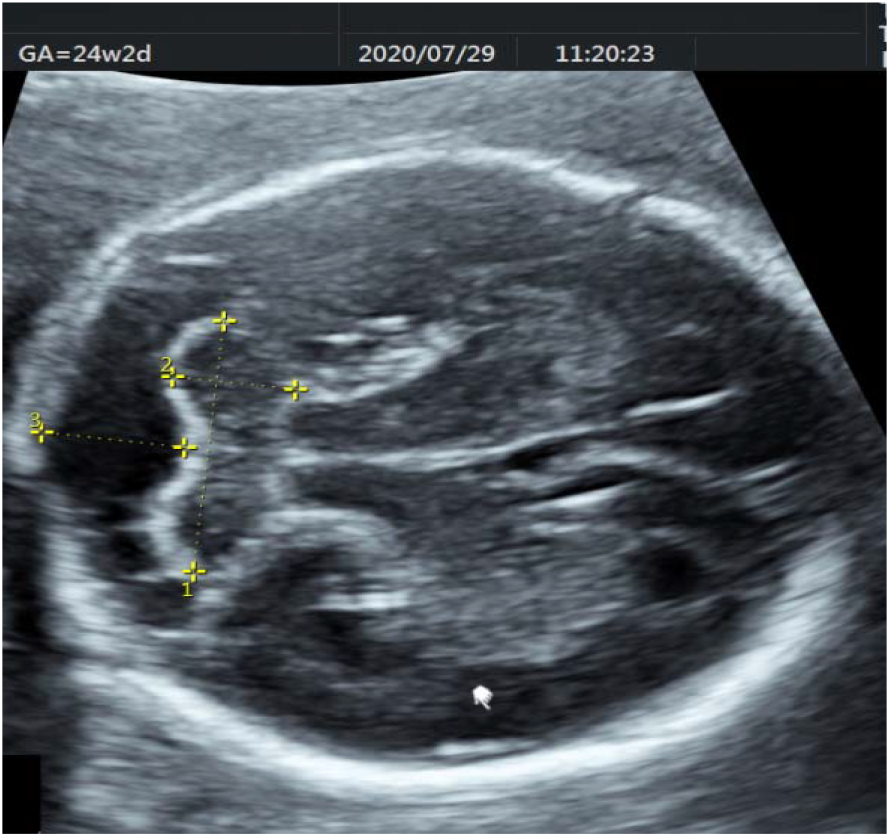
fetus diagnosed microcephaly with Sylvian fissure absence at 24 weeks of gestation

**Fig. 5.**
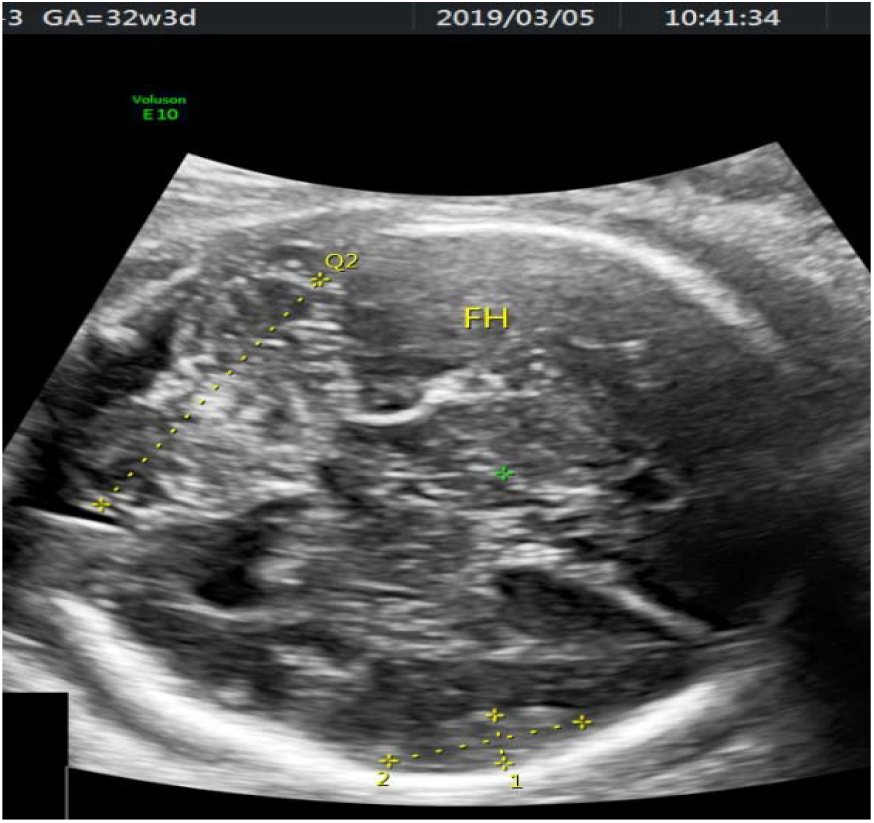
fetus diagnosed aplasia of corpus callosum at 32 weeks of gestation with the angle of posterosuperior horn of the Sylvian fissure above the 95th percentile and the still shown 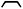 shape like 21 ∼ 24 weeks)

In this study, we innovatively selected trans-cerebellar section to evaluate the Sylvian fissure of normal fetuses and preliminarily elucidated the angle of posterosuperior horn of the Sylvian fissure of normal fetuses at 21 to 32 weeks of gestation and its morphological changes, which can provide valuable information for the evaluation of the normal development of fetal Sylvian fissure and the prenatal diagnosis of fetal cortical dysplasia.

## Data Availability

Data availability. The full data that supports the findings of this study are available from the corresponding author upon reasonable request.

## ACKNOWLEDGEMENTS

The authors thank all the women who participated in the study and acknowledge their significant contribution. Thank you to Zhang Dirong and Shi Yu for helping to design the research methods, Kong Fengbei and Yao Chunxiao for support with statistical analysis, She Ying and Wu Guoru for proposing some strategies for writing.

